# Estimating the Genetic Contribution to Astigmatism and Myopia in the Mexican population

**DOI:** 10.1101/2023.02.18.23286098

**Authors:** Talía V. Román-López, Brisa García-Vilchis, Vanessa Murillo-Lechuga, Enrique Chiu-Han, López-Camaño Xanat, Oscar Aldana-Assad, Santiago Diaz-Torres, Ulises Caballero-Sánchez, I. Ivett E. Ortega-Mora, Diego Ramirez-Gonzalez, Diego Zenteno, Zaida Espinosa-Valdés, Andrea Tapia-Atilano, Sofía Pradel-Jiménez, Miguel E. Rentería, Alejandra Medina-Rivera, Alejandra E. Ruiz-Contreras, Sarael Alcauter

## Abstract

Astigmatism and myopia are two common ocular refractive errors that can impact daily life, including student learning and productivity. Current understanding suggests that the etiology of these conditions is the result of a complex interplay between genetic and environmental factors. Studies in populations of European ancestry have demonstrated a higher concordance of refractive errors in monozygotic (MZ) twins compared to dizygotic (DZ) twins. However, there is a lack of studies on genetically informative samples of multi-ethnic ancestry. This study aimed to estimate the genetic contribution to astigmatism and myopia in the Mexican population. A sample of 1399 families, including 243 twin pairs and 1156 single twins, completed a medical questionnaire about their own and their co-twins diagnosis of astigmatism and myopia. Concordance rates for astigmatism and myopia were estimated in Mexican twins, and heritability and genetic correlations were determined using a bivariate ACE Cholesky decomposition method. The results showed a concordance rate of 0.74 for MZ twins and 0.50 for DZ twins for astigmatism, and a higher concordance rate in MZ twins, 0.74, compared to DZ twins, 0.54 for myopia. Heritability estimates were 0.66 for astigmatism and 0.62 for myopia, with a cross-trait genetic correlation of rA=0.82 and a phenotypic correlation of rP=0.80. These results are consistent with previous findings in other populations, providing evidence for a similar genetic architecture of these conditions in the multi-ethnic Mexican population.

## Introduction

Astigmatism and myopia are two prevalent ocular refractive errors that have become significant public health concerns globally (Baird et al., 2020; Hashemi et al., 2017; Pascolini & Mariotti, 2012). Astigmatism is characterized by unequal curvatures in the cornea or crystalline lens, leading to rotational asymmetries and blurry projections of light over the retina (Harb & Wildsoet, 2019; Harris, 2000; Visnjić et al., 2012). On the other hand, myopia, also known as nearsightedness, is caused by the light being focused in front of the retina instead of on it, leading to the blurred perception of distant objects (Harb & Wildsoet, 2019). The elongation of the eye and corneal modifications (*e*.*g*., keratoconus) can contribute to myopia (Baird et al., 2020).

The worldwide prevalence of myopia was estimated to be ∼33% by the World Health Organization in 2020, and a meta-analysis of global studies estimated a prevalence of 26.5% for myopia and 40.4% for astigmatism (Holden et al., 2016). However, data varies greatly between regions and ethnic groups, with higher prevalence in some groups (Hashemi et al., 2017; Rose et al., 2001), for example, in East and Southeast Asia, myopia is considered an epidemic among adults, with 80-90% suffering from it (Morgan et al., 2018); half of the european population sufffer some refraction error, around 30% of myopia and 23% of astigmatism (Williams et al., 2015). Meanwhile, data in other regions, such as Latin America, is scarce. In Mexico, astigmatism and myopia are common ocular problems (Secretaría de Salud, 2020). For instance, in a sample of 676,856 Mexican patients (aged 6 to 90), myopia was the most common refractive error at 24.8%, while astigmatism was 13.5% (Gomez-Salazar et al., 2017). Studies of school-age children in urban areas showed 44% bilateral myopia and 9.5% astigmatism, while those in rural areas showed 9.7% and 4.4% respectively (Garcia-Lievanos et al., 2016). Refractive errors impact aspects of life such as education and employment (Kandel et al., 2017). The concern about these conditions is growing, as it is predicted to affect over 50% of the world’s population by 2050 (Holden et al., 2016), thus, evaluating the etiology of refractive errors is crucial.

Previous research suggests that both genetic and environmental factors play a role in the development of astigmatism and myopia (Baird et al., 2020; Gordon-Shaag et al., 2021; Read et al., 2007; Young et al., 2007). For instance, genome-wide association studies (GWAS) have identified various risk polymorphisms for both conditions, including genes involved in eye growth, retina proteins, corneal epithelium, neurotransmission, and retinoic acid metabolism (Harb & Wildsoet, 2019; Hysi et al., 2010; Kiefer et al., 2013; Lopes et al., 2013; Nakanishi et al., 2009; Shah, Guggenheim, et al., 2018; Wojciechowski, 2011; Wojciechowski & Hysi, 2013). However, the genetic connection between astigmatism and myopia remains inconclusive, with some studies suggesting a shared genetic aetiology and others considering them as different manifestations of refractive errors (Pinazo-Durán et al., 2016; Shah, Guggenheim, et al., 2018; Young et al., 2007; Dirani et al., 2008; Hammond et al., 2001; Paget et al., 2008). Environmental factors, such as prolonged near-work activities, outdoor time, reduced sleep, education, muscle changes, and population density, also play a role (Demir et al., 2021; Harb & Wildsoet, 2019; Li et al., 2019; Saad & El Bayoumy, 2007; Wang et al., 2021; Wojciechowski, 2011; Xiong et al., 2017; Zhang et al., 2010).

Twin studies are useful in evaluating the combined impact of genetics and environment (Sahu & Prasuna, 2016). For example, a study in Norway showed higher concordance rates of astigmatism in monozygotic twins than dizygotic twins, suggesting a greater genetic influence (Grjibovski et al., 2006). The heritability of astigmatism was estimated to be over 60% in an Australian twin study (Dirani et al., 2008). A Chinese twin study also found significant contributions from both genetics and environment to myopia (Chen et al., 1985). Despite these studies, it is needed to evaluate the contribution of genetics and environment in genetically diverse populations, such as Mexicans, which have been underrepresented in genetic studies and where refractive errors have a high prevalence. This study aims to determine the concordance rates, heritability, and genetic cross-trait correlation of astigmatism and myopia in Mexican twins.

## Methods

### Sample

Data used for this study comes from the Mexican Twin Registry, TwinsMX (https://twinsmxofficial.unam.mx/; Leon-Apodaca et al., 2019), collected using the Research Electronic Data Capture (REDCap) platform, hosted at the National Laboratory of Advanced Scientific Visualization at the Universidad Nacional Autónoma de México (UNAM). All participants gave informed consent and the study protocol was reviewed and approved by the Research Ethics Committee of the Institute of Neurobiology at UNAM. All data is stored and managed in compliance with the “Mexican Federal Law on the Protection of Personal Data Held by Private Parties”.

At the time of data extraction (April 2022), TwinsMX included data for 2,778 families. For this study, we selected subjects who completed the medical questionnaire and were aged seven years or older (considering the start of scholar-age can vary between 6-7 years old in Mexico), resulting in a sample of *n*=1887 families. Zygosity status was participant-reported, twin pairs whose reported zygosity did not match (*e*.*g*., one twin reported MZ and the co-twin reported DZ) were classified as indeterminate and were excluded (Sánchez-Romera, 2013). Also, subjects from other multiple birth types (e.g., triplets or cuadruplets) were excluded. The final sample consisted of *n*=1,399 families. A family was defined as either singleton or a pair of twins. In this study, 243 families with both twins being registered (*i*.*e*. 486 individuals) and 1,156 families with only one registered twin were included in the final sample. The 1,156 single twins reported information about their unregistered twin, with this information we are able to analyze a sample of *n*=2,798 individuals (*i*.*e*., 486 + (1156*2)). Sociodemographic data about sex and age of the twins were also acquired.

### Myopia and astigmatism participant-reported diagnosis

Twins answered a medical questionnaire where they were asked, “*Have you, your parents, siblings, or children ever suffered some of the following conditions?”*, and tick boxes allow participants to state which family members present the condition. Among the answers were enlisted myopia and astigmatism.

### Statistical Analyses

Participants were split into two main groups: All MZ and All DZ, based on the self reported zygosity. Additionally, each twin reported their sex and their twin sex. With that information, families were classified into five different subgroups depending on zygosity and sex (MZF, MZM, DZF, DZM, and DZOs), as has been widely reported (*e*.*g*. Grjibovski et al., 2006; Hopper et al., 1990; Loat et al., 2004; Vink & Boomsma, 2011).

The participant-report diagnosis was used for families where both twins were part of the registry. For the families where only one of the twins was part of the registry (single twins), it was considered the report about themselves and the report about their twin.

To address the concern of reliability of the twin reported diagnosis when only one of the twins was available, we adopted the following strategy: first, we analyzed the responses from the 243 twin pairs (both twins registered) and tested the consistency of their answers regarding their co-twin. Additionally, it was estimated the concordance rate for the diagnoses of the 243 twin pairs to compare it with that obtained with the whole sample.

#### Demographic analysis

We compared the distribution of sex and age between the MZ and DZ groups using an independent chi-square test (χ2).

#### Concordance rate test

We calculated probandwise concordance for both astigmatism and myopia following the model reported by McGue (1992), and calculated the respective confidence intervals for proportions for each group and subgroups of zygosity and sex. Due to the small sample size of some of the zygosity and sex subgroups, only the comparisons between concordance rates for All MZ and All DZ groups, without stratification by sex, were tested with the Likelihood Ratio Test. Only results with *p* < 0.05 were considered statistically significant.

#### Bivariate ACE Cholesky analysis

We performed the ACE Cholesky decomposition, which allows estimating the amount of the variance of each phenotype, explained by the genetic contribution or heritability (A), the shared environmental contribution (C), and the unique environment (E). In addition, a multivariate design (in this case, a bivariate model) allows for estimating the covariation between myopia and astigmatism. For a detailed description of these analyses, see Zietsch et al. (2014) and Posthuma (2009).

Briefly, the bivariate model assumes that latent variables are having effects on the traits of interest (see Figure 1 for the path model). First, considering the genetic contribution from two sets of genes (latent variables A1 and A2) by associating directly first gene set over one trait (*i*.*e*. A1 over astigmatism) through a path (a_11_), and the second set of genes contributing over the second trait (*i*.*e*. A2 acting over myopia) through a second path (a_22_). Second, the model takes into account the genes shared between astigmatism and myopia, which are modeled in the influence of A1 over the second trait, myopia, through path a_21_. The effect of the set of genes A2 over the first trait (astigmatism) via a_12_ is not modeled to avoid redundancy; namely, it is assumed that if an overlapping of shared genes exists, these will be the same group of genes within A1 or A2 sets, then the path a_21_ is already reflecting the conjunction of shared genes. Additionally, it is relevant to notice that, in a Cholesky factorization, the lower triangular solution is mathematically equivalent to the upper triangular solution (*see* matrix *a* below). On the other hand, the respective shared and nonshared environmental contributions are correspondingly modeled by C and E from paths.

**Figure 1.**
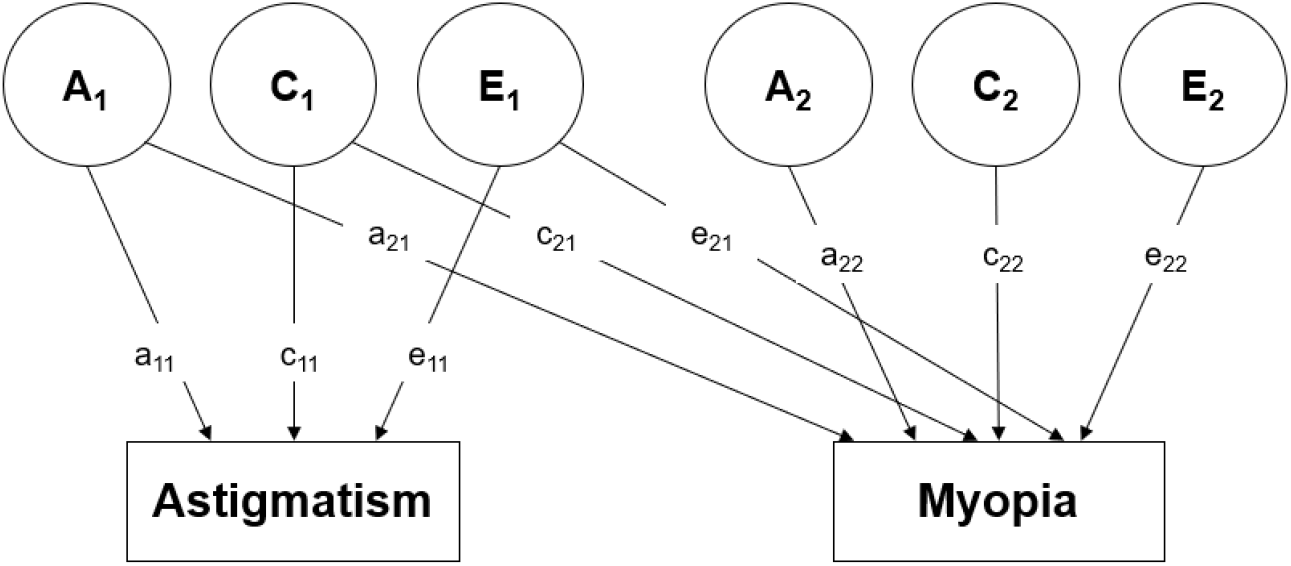
Bivariate path modelling for astigmatism and myopia. Cholesky decomposition in latent variables: A (genetic contribution), C (shared environment influence), and E (residual or non-shared environmental influences). A1 represents the latent variable (*i*.*e*. the set of genes) that contributes to astigmatism (path a_11_) and myopia (path a_21_). A2 is the second latent variable (i.e. a second set of genes) affecting myopia. As well as the respective variables for shared and nonshared environmental contributions (C and E).

The corresponding matrix design of this bivariate path model is an *n* x *n* matrix, where *n* is the number of traits in the model, in this case, a 2 × 2 matrix 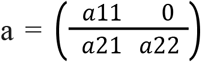. The total genetic contribution is estimated as the result of A=a*a^T^, given as a result 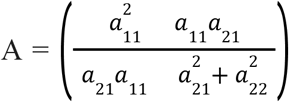. The A(1,1) = a_11_^2^ is the total genetic contribution (*i*.*e*., heritability) of the trait 1, A(2,2) = a_21_^2^ + a_22_^2^, is the total genetic contribution (*i*.*e*. heritability) of the trait 2. Meanwhile, the cross-trait cross-twin genetic covariance is A(2,1) = a_11_a_21_. To estimate the genetic correlation between the traits of interest, astigmatism and myopia, 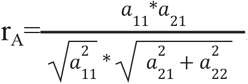. The variance and covariance matrices, and correlations for C and E can be calculated analogously.

Data analysis was performed in Ubuntu 22.04 using RStudio v.4.2.0 (2022-04-22, SCR_000432) and packages: tidyverse (v.1.3.1, Wickham et al., 2019, SCR_019186), gt (v.0.6.0, Iannone et al., 2022) and the UMX package, (v.4.10.50 and OpenMx v2.20.6) (Bates et al., 2019) were used for the bivariate structural modeling of the ACE Cholesky decomposition. For the umxACE function, the arguments addCI and Intervals were set up as True, the modeling was performed using the “CSOLNP” optimiser. All code is available in GitHub URL: https://github.com/NeuroGenomicsMX/TwinsMX_Astigmatism_Myopia.

## Results

Considering that DZ twin pairs can be discordant for sex, we performed a chi-square test between MZ and DZ for the total sample by sex. The chi-square test did not show significant differences in sex ratios between DZ and MZ (χ^*2*^(1, N=2798) = 1.83, *p*=0.18). Figure 2 shows subgroups or pairs segregated by zygosity and sex (2A) and distribution by age group (2B). No differences in distribution by age group were observed between MZ and DZ pairs (χ^*2*^(4, N=1399) = 7.1623, *p*=0.13).

**Figure 2.**
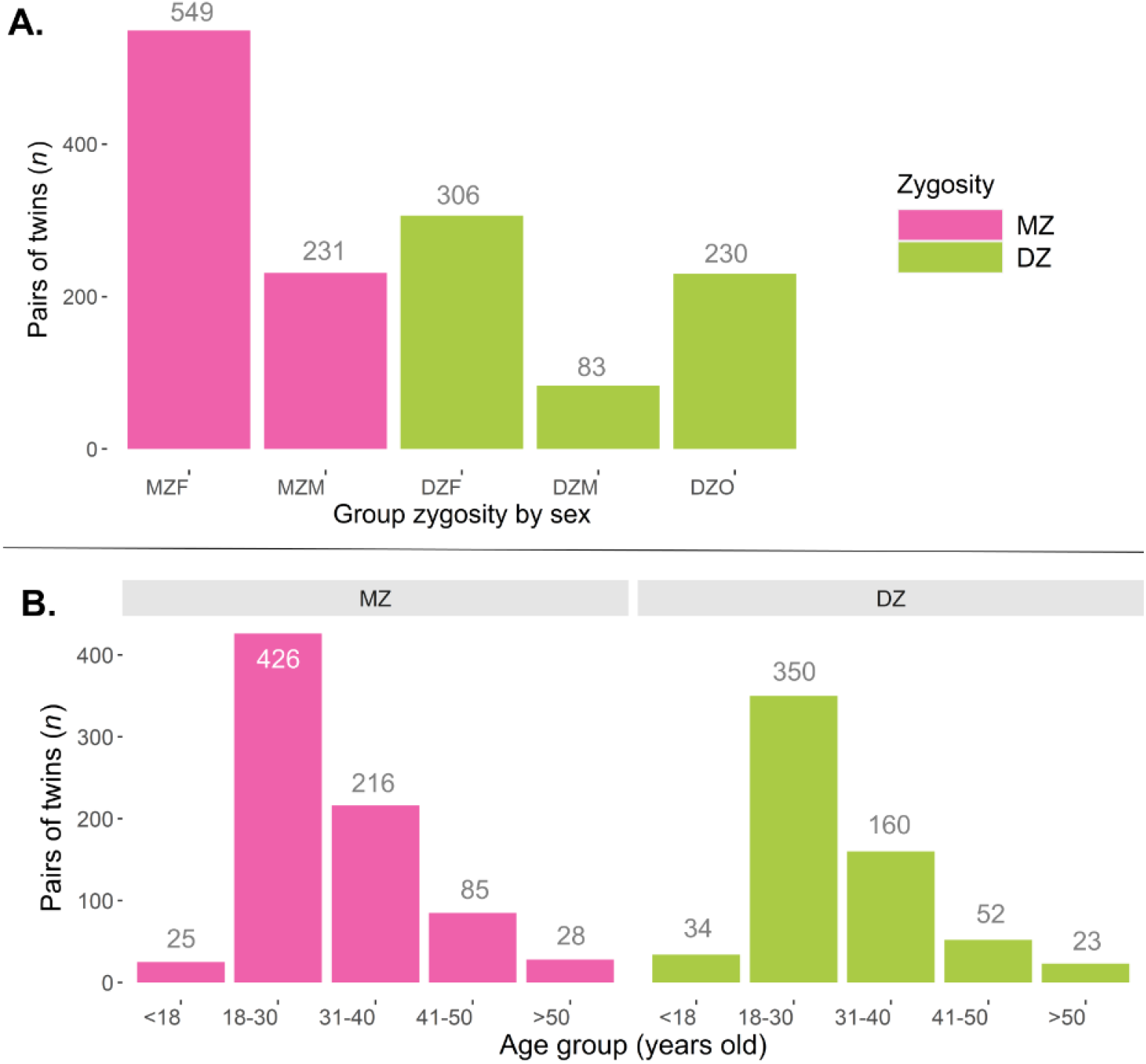
**A. Distribution of pairs segregated by zygosity and sex**. Sample size by group no differences by group were observed (*p*=0.18). **B. Distribution of pairs segregated by zygosity and age group**. No differences between DZ and MZ pairs were observed (*p*=0.13).

### Concordance Rates

#### Astigmatism

Among 1399 families, the incidence of astigmatism was 36% (1016/2798). Concordance rates results showed that MZ twins had a significantly higher astigmatism concordance than DZ (χ^2^(1)= 40.20, *p*=2.29×10^−10^).

#### Myopia

The incidence of myopia was 45% (1269/2798). The concordance rate was significantly higher for MZ than for DZ twins (χ^2^(1)= 33.09, *p*=8.80×10^−9^).

### Additional analysis for complete pairs only

The same statistics were estimated for the subsample that included the participant-report of both twins (243 pairs) for both MZ and DZ groups. Consistent with the previous results (considering the report from one twin for both twins), MZ twins showed higher concordance rates for astigmatism (χ^2^(1)= 14.72, *p*=1.20×10^−4^; Table 3) and myopia (χ^2^(1)= 12.08, *p*=5.0×10^−4^; Table 4).

**Table 1.** Astigmatism concordance rates in Mexican twin pairs.

| Group |  | Total<br>*n* | Pairs | Positive<br>Cases *n* | Negative<br>Cases *n* | Concordant<br>pairs | Discordant<br>pairs | Probandwise<br>concordance<br>rate | 95% CI |
| --- | --- | --- | --- | --- | --- | --- | --- | --- | --- |
| **MZ** | MZF | 1098 | 549 | 439 | 659 | 165 | 109 | 0.75 | 0.71-0.79 |
|  | MZM | 462 | 231 | 138 | 324 | 49 | 40 | 0.71 | 0.63-0.79 |
| **DZ** | DZF | 612 | 306 | 249 | 363 | 74 | 101 | 0.59 | 0.53-0.66 |
|  | DZM | 166 | 83 | 40 | 126 | 11 | 18 | 0.55 | 0.40-0.70 |
|  | DZOs | 460 | 230 | 150 | 310 | 36 | 78 | 0.48 | 0.40-0.56 |
| All MZ |  | 1560 | 780 | 577 | 983 | 214 | 149 | 0.74 | 0.71-0.78 |
| All DZ |  | 1238 | 619 | 439 | 799 | 121 | 197 | 0.50 | 0.50-0.60 |
| **Total** |  | 2798 | 1399 | 1016 | 1782 | 335 | 346 | 0.66 | 0.63-0.69 |

$\chi^2(1) = 40.20$ , $p = 2.29 \times 10^{-10}$

**Table 2.** Myopia concordance rates in Mexican twins pairs.

| Group |  | Total<br>*n* | Pairs | Positive<br>Cases *n* | Negative<br>Cases *n* | Concordant<br>pairs | Discordant<br>pairs | Probandwise<br>concordance<br>rate | 95% CI |
| --- | --- | --- | --- | --- | --- | --- | --- | --- | --- |
| **MZ** | MZF | 1098 | 549 | 538 | 560 | 209 | 120 | 0.69 | 0.64-0.74 |
|  | MZM | 462 | 231 | 165 | 297 | 65 | 35 | 0.47 | 0.33-0.61 |
| **DZ** | DZF | 612 | 306 | 321 | 291 | 111 | 99 | 0.58 | 0.51-0.65 |
|  | DZM | 166 | 83 | 51 | 115 | 12 | 27 | 0.78 | 0.74-0.81 |
|  | DZOs | 460 | 230 | 194 | 266 | 56 | 82 | 0.79 | 0.73-0.85 |
| All MZ |  | 1560 | 780 | 703 | 857 | 274 | 155 | 0.74 | 0.71-0.78 |
| All DZ |  | 1238 | 619 | 566 | 672 | 179 | 208 | 0.55 | 0.50-0.60 |
| **Total** |  | 2798 | 1399 | 1269 | 1529 | 453 | 363 | 0.66 | 0.63-0.69 |

**Table 3.** Astigmatism concordance rates in Mexican twin pairs.

| Group |  | Total<br>*n* | Pairs | Positive<br>Cases *n* | Negative<br>Cases *n* | Concordant<br>pairs | Discordant<br>pairs | Probandwise<br>concordance<br>rate | 95% CI |
| --- | --- | --- | --- | --- | --- | --- | --- | --- | --- |
| All MZ |  | 324 | 162 | 151 | 173 | 55 | 41 | 0.73 | 0.66-0.80 |
| All DZ |  | 162 | 81 | 66 | 96 | 15 | 36 | 0.45 | 0.33-0.57 |
| **Total** |  | 486 | 243 | 217 | 269 | 70 | 77 | 0.65 | 0.58-0.71 |

**Table 4.**
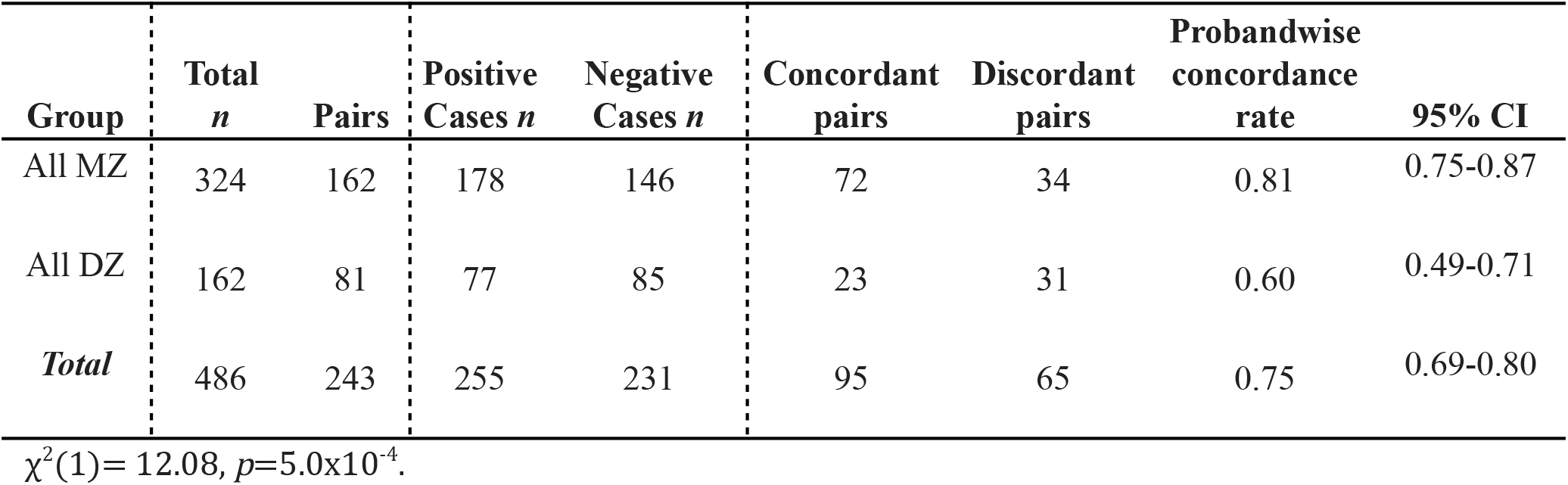
Astigmatism concordance rate in Pairs of Mexican Twins.

| **Group** | **Total**<br>***n*** | **Pairs** | **Positive**<br>**Cases *n*** | **Negative**<br>**Cases *n*** | **Concordant**<br>**pairs** | **Discordant**<br>**pairs** | **Probandwise**<br>**concordance**<br>**rate** | **95% CI** |
| --- | --- | --- | --- | --- | --- | --- | --- | --- |
| All MZ | 324 | 162 | 178 | 146 | 72 | 34 | 0.81 | 0.75-0.87 |
| All DZ | 162 | 81 | 77 | 85 | 23 | 31 | 0.60 | 0.49-0.71 |
| **Total** | 486 | 243 | 255 | 231 | 95 | 65 | 0.75 | 0.69-0.80 |

$\chi^2(1) = 12.08, p = 5.0 \times 10^{-4}$ .

### Heritability and cross-trait correlation

The heritability (A) of astigmatism was estimated to be 0.66, with shared environmental effects contributing 0.14 (C) and residual or non-shared environmental contributions of 0.20 (E). Meanwhile, heritability for myopia was 0.62 (A), with C = 0.18 and E = 0.20. Values of each estimate in the model (*i*.*e*. a_11_, a_21_, …) and their corresponding 95% CI are reported in Table 5.

**Table 5.** Estimation for each parameter in bivariate modeling’ and 95% CI.

|  | **Estimate** | **95% CI** |
| --- | --- | --- |
| $a_{11}$ | 0.81 | (0.76-0.83) |
| $a_{21}$ | 0.64 | (0.47-0.79) |
| $a_{22}$ | 0.45 | (0.37-0.56) |
| $c_{11}$ | 0.37 | (0.06-0.46) |
| $c_{21}$ | 0.31 | (-0.07-0.59) |
| $c_{22}$ | 0.29 | (0.00001-0.37) |
| $e_{11}$ | 0.45 | (0.43-0.51) |
| $e_{21}$ | 0.40 | (0.36-0.46) |
| $e_{22}$ | 0.20 | (0.16-0.21) |

Additionally, bivariate modeling allowed to estimate the cross-trait correlation, for this model the genetic correlation was r_A_=0.82, with the shared environmental correlation r_C_=0.73, and non-shared environmental correlation r_E_ = 0.89. Finally, the phenotypic correlation between astigmatism and myopia was r_P_ = 0.80.

## Discussion

This study aimed to estimate the concordance rates and heritability of myopia and astigmatism in Mexican twins. Given the genetically diverse ancestral composition of the Mexican population (García-Ortiz et al., 2021; Martínez-Cortés et al., 2012) this is the first study to provide these results specifically for a Mexican twin sample. The results showed higher concordance rates for myopia and astigmatism in monozygotic (MZ) twins compared to dizygotic (DZ) twins. The estimated heritability was 0.66 for astigmatism and 0.62 for myopia, and the genetic correlation (rA=0.82) suggests that both traits are influenced by a shared set of genes. Although a correlation lower than one does not necessarily imply that the set of shared genes has a similar effect on both astigmatism and myopia (Posthuma, 2009), the high value of the genetic correlation in this study supports that astigmatism and myopia share a genetic basis and overlap in their genetic effects.

A genome-wide association study (GWAS) in an European ancestry sample found that the NPLOC4/TSPAN10 (17q25.3) gene cluster, which has previously been linked to myopia and other ocular disturbances (*e*.*g*. Plotnikov et al., 2019), was also associated with astigmatism (Shah, Li, et al., 2018). Another study in individuals from the UK and Canada showed that keratoconus, a corneal deformity and thickness associated with early stages of myopia and astigmatism, involved approximately 500 genetic loci, suggesting a highly polygenic architecture of ocular refraction errors (He et al., 2022). The present study highlights the genetic overlap between astigmatism and myopia, and further research is needed to identify the specific shared or unique loci contributing to the etiology of these conditions. The higher concordance rates and heritability estimates in this study indicate that these refractive errors have a strong genetic contribution in the Mexican population.

Our results are consistent with prior research on the heritability of astigmatism and myopia in various populations. For astigmatism, studies have demonstrated greater heritability by observing higher correlations within MZ twins compared to DZ twins for factors such as refractive error, axial length, and corneal curvature (Dirani et al., 2008; Lyhne et al., 2001; Teikari et al., 1989). In the case of myopia, a Chinese study from 1987 found a higher concordance rate in MZ twins (0.65) than in DZ twins (0.46) (Lin & Chen, 1987). Our findings, with concordance rates of 0.74 and 0.55 for MZ and DZ twins, respectively, reveal a similar trend. Currently, it is understood that myopia results from the interplay of multiple genes and genetic variants that influence eye growth and retinal signaling (Williams et al., 2017).

The demographic analyses showed no differences in the distribution of age group nor sex between MZ and DZ twins, suggesting that differences in demographics (*p*>0.05) do not explain our results. Additionally, our results show a higher prevalence of myopia (45%) than astigmatism (36%) in the Mexican population, which is consistent with those previously observed by Gomez-Salazar et al. (2017).

On the other hand, while biometric measures are widely recognized as a more reliable method to diagnose astigmatism (Diranis et al., 2008), their use can limit the extent of participant recruitment, particularly in populations like Mexico, where obtaining large sample sizes with biometric measures is a geographic and economic challenge. In these circumstances, participant self-reported data acquired through online methods can offer a significant advantage for twin studies, particularly in terms of size and geographic representation (Grjibovski et al., 2006; Hur et al., 2019).

Our results were robust even when considering reports from only one of the twins. The primary analysis conducted on 1399 families and the analysis on 243 complete pairs both replicated the results for myopia and astigmatism. Furthermore, the consistency of participant and co-twin reports was observed to be high, with 80.45% agreement for astigmatism and 84.36% for myopia. This suggests that the participant and co-twin reports were highly reliable, and supports the value of using participant-reported data in twins studies, especially when only one of the twins can provide information. This method allows the effective use of data obtained through electronic records, making research possible for underrepresented populations. Nevertheless, further research should compare clinical diagnoses and participant-reported data to assess the similarity of results and address this inherent limitation in participant-reported data.

One shortcoming of the study is that the limited sample size prevented us from conducting subgroup analyses by zygosity and sex. Future research should aim to overcome this limitation by increasing the sample size, in order to investigate genetic differences as a function of sex in greater detail. Given the high genetic influence demonstrated in the current results, it is also desirable to explore possible genetic factors and variations in the Mexican population through techniques such as Genome-Wide Association Studies (GWAS) (Nakanishi et al., 2009; Shah et al., 2018). Despite the strong genetic component, it is also important to consider environmental factors such as lifestyle, nutrition, the use of electronic devices, and near-work that may play a role in the prevalence of myopia and astigmatism in the Mexican population.

It is not unexpected that one of the first twin studies on the topic focused on examining the concordance of refraction in human eyes. Twin studies afford a unique chance to investigate conditions like astigmatism and myopia. In conclusion our study affirms that the likelihood of developing astigmatism and myopia in the Mexican population is significantly shaped by genetic factors.

## Data Availability

All data produced in the present study will be available online after being published.

## Acknowledgments

We would like to thank all the twins for their willingness to participate and for taking the time to be part of the registry. To the social media and designers team: Mauricio Guzman and Andrea Bermeo. And to all the team of the TwinsMX: Regina Casa Madrid, Ian Espinosa, Arantza Piña, Brenda Gónzalez, Xochil Díaz, and Itzamná Sanchez for their support in twin experimental sessions. This work received technical support from Luis Aguilar, Alejandro León, and Jair García of the Laboratorio Nacional de Visualización Científica Avanzada. We also thank Carina Uribe Díaz, Leopoldo González Santos and Alejandra Castillo Carbajal for their technical support.

## Financial support

This work was supported by the Consejo Nacional de Ciencia y Tecnología (CONACYT) (Grant CF-2019 No. 6390). In addition, MER is supported by the Al & Val Rosenstrauss Fellowship from the Rebecca L. Cooper Medical Research Foundation, Australia (F20231230).

## Conflict of interest

None.

## Ethical standards

*The authors assert that all procedures contributing to this work comply with the ethical standards of the relevant national and institutional committees on human experimentation and with the Helsinki Declaration of 1975, as revised in 2008*.

## Notes

### Competing Interest Statement

The authors have declared no competing interest.

### Funding Statement

This study was funded by the Consejo Nacional de Ciencia y Tecnologia (CONACYT) (Grant CF-2019 No. 6390). In addition, MER is supported by the Al & Val Rosenstrauss Fellowship from the Rebecca L. Cooper Medical Research Foundation, Australia (F20231230).

### Author Declarations

Ethics Committee of the Institute of Neurobiology at UNAM gave ethical approval

